# Predicting Tuberculosis Incidence in Adult HIV Patients on ART in Debre Markos, Ethiopia: A Machine Learning Approach

**DOI:** 10.1101/2025.08.24.25334330

**Authors:** Desalegn Meseret Tadele, Getaye Tizazu Biwota, Lijalem Megibaru Enyew, Maru Meseret Tadele, Gizaw Hailiye Teferi

## Abstract

Tuberculosis (TB) is the commonest comorbidity among individuals with HIV/AIDS, especially in low- and middle-income nations such as Ethiopia. Early diagnosis of TB infection in HIV-infected patients is crucial for effective management of opportunistic infections that can result in mortality. Early identification of TB in HIV/AIDS patients plays a significant role in reducing morbidity and mortality. Machine learning algorithms have a significant role in detecting TB occurrences among HIV/AIDS patients.

In this study, we used 5,392 HIV-infected individuals’ medical records. Techniques such as SMOTE and ADASYN were employed to adjust data imbalance between positive and negative TB status. Random forest, decision tree, logistic regression, gradient boosting, K-nearest neighbors, and XGBoost were evaluated to predict TB incidence.

Of the total records, 3,440 (63.8%) were female patients, and the remaining 1,952 (36.2%) were male patients. 3,715 (68.9%) were labeled as green records addresses, while 1,677 (31.1%) had yellow records. The XGBoost algorithm is the best-performing model to predict TB incidence. Among the features included in this study is the most important classifier for feature selection. Among all the features, CD4 count and patient age were found to be the most important predictors of TB incidence among adult HIV patients.

This study demonstrates that the XGBoost model was the most effective model for predicting tuberculosis incidence among HIV patients, utilizing features such as low CD4 counts, age, duration on ART, weight, sex, and WHO clinical stage.

**Author Summary:** Tuberculosis is the main opportunistic infection among HIV-infected individuals. The comorbidities of HIV and TB increase the risk of mortality among HIV patients. Early detection of TB infection from people living with HIV is a crucial step for increasing HIV patients’ life expectancy. A machine learning approach plays a vital role in detecting TB infection among HIV patients. This study aims to predict tuberculosis (TB) occurrence in adult HIV patients on antiretroviral medication (ART) in Debre Markos City, Ethiopia. Using a retrospective dataset of 5,392 patients, researchers compared seven methods. The XGBoost model fared best, earning 82% accuracy and a 90% AUC after resolving class imbalance using MOTE+ENN. Key predictors revealed were low CD4 count, age, time on ART, sex, WHO clinical stage, address status, DSD category, and TB preventative treatment (TPT status). Machine learning models will help health providers to predict the risk of TB infection and make early intervention in high TB-HIV co-infection loads.

## Introduction

HIV is an infectious virus that primarily undermines the immune system by infecting specifically the CD4 cells, or T cells, which are responsible for combating various infections. The weakening of the body’s defense system through this process predisposes one to opportunistic diseases (OIs), such as tuberculosis [1]. Acquired Immune Deficiency Syndrome (AIDS) is a condition that occurs secondary to HIV infection. Tuberculosis (TB) is one of the common opportunistic infections that affect people with HIV/AIDS when compared with non-HIV-infected individuals [2, 3].

Tuberculosis is the primary opportunistic illness in individuals infected with HIV, and the likelihood of developing active tuberculosis markedly increases owing to the HIV virus. HIV compromises the immune system, increasing the likelihood of tuberculosis infection advancing from latent to active phases, hence rendering co-infected patients more susceptible to mortality [4].

Globally, HIV-positive adults are highly susceptible to TB, and an estimated 208,000 ALWHIV died from TB in 2020. Sub-Saharan Africa, including Ethiopia, experiences high co-infection rates due to the synergy between HIV and TB, where declining immunity from HIV increases the risk of developing active TB [5].

Several studies locally have been conducted to determine the incidence and predictors of TB among adult HIV patients using classical statistical methods. Which primarily focus on learning factors behind TB cases that already exist. While they identify general risk factors, they do not offer personalized risk assessments. Additionally, they are static in nature because any patient data or new information does not update the analysis automatically. Moreover, the results are not as easily translated into real-time clinical applications.

To overcome these challenges, this study used machine learning (ML) models to predict the incidence and identify the predictors of TB among adult HIV patients. ML emerged as a promising approach for analyzing complex health data and forecasting disease incidence [6]. Debre Markos City hosts numerous public health facilities providing ART services to adult HIV patients. Even though the facilities in the city have a strong electronic health record, none of them used the historical data to develop a model for predicting the risk of incidence of TB prior to the occurrence of the event. Therefore, the aim of this research is to develop a model for predicting the risk of TB in adult HIV patients on ART in public health facilities in Debre Markos City using past data. Therefore, the aim of this research is to develop a model for predicting the risk of TB in adult HIV patients on ART in public health facilities in Debre Markos city using past data.

Adults on ART who develop TB may also have more advanced disease and complications, including respiratory distress and systemic illness, due to the dual burden of infection. Mortality risk was considerably high in TB patients who were left untreated compared to those who did not have TB [7].

HIV-TB co-infection considerably diminishes the health-related quality of life in adult HIV patients in the areas of physical health, mental health, and social functioning. Both diseases can cause stigma, which may result in social isolation and mental illness. The onset of TB may disrupt adherence to antiretroviral treatment (ART) because of the complexities of managing many drugs and the side effects of TB treatment [8].

Active TB patients are still infectious while on ART, which makes it even more community transmitted. In high HIV prevalence settings, the co-occurrence of TB can cause outbreaks, thus making it a public health issue. It requires a lot of healthcare services to treat dual-diagnosed patients, including longer hospital stays, additional diagnostics, and specialized care teams.

TB development on ART patients presents significant challenges, both to individual health outcomes and to public health efforts at large. Addressing these issues requires coordinated healthcare strategies, ongoing research, and comprehensive support systems for affected individuals. This study aims to predict the incidence of TB among adult HIV patients and evaluate which model predicts accurately. Machine learning has emerged as a powerful force in healthcare as a predictor of disease outcomes. The innovation in machine learning and data analytics within the healthcare sector creates new possibilities in the optimization of patient care. Machine learning algorithms can process intricate data sets, thereby detecting patterns and connections that standard statistical techniques may fail to capture. Through the integration of various data sources and the detection of complicated patterns, the algorithms provide a robust alternative that results in higher predictive power. Hence, these innovative methods can facilitate a more precise targeting of individuals in adult HIV cohorts who are at risk of developing tuberculosis [9].

Capacity to process big and intricate datasets enables detection of non-linear relationships and interdependencies between variables, which might not be caught using conventional statistical techniques. Machine learning in medicine has gained speed; most recently, for disease prediction, it demonstrated the potential of machine learning models for predicting TB in adult HIV patients, indicating that these technologies can enhance clinical decision-making [10]. Machine learning models can process big datasets to identify patterns that are not visible to traditional approaches, representing a promising avenue for patient outcome enhancement. Ethiopia is one of the low-income countries that is heavily affected by both tuberculosis (TB) and HIV, with the country featuring in the top 30 of the world’s TB incidence rate, in addition to a high coincidence of TB and HIV cases [11]. Within this framework, the Debre Markos health facility functions as an essential health provider, addressing the needs of at-risk populations for these conditions. Researchers have revealed that there are significant challenges in the management of patients with co-infections, ranging from restrictions on resources, inadequate screening protocols, and limitations on data collection. The literature demands that there is a pressing need to tackle the TB and HIV co-infection with special reference to resource-poor settings such as Ethiopia [12]. The application of machine learning techniques is a promising way forward to add to predictive effectiveness and optimize patient care [13].

## Methods

### Study design, setting, and study population

An institution-based retrospective cross-sectional study design was employed at Debre Markos Comprehensive Specialized Hospital and Debre Markos Health Center in Debre Markos City, Ethiopia.

Records of all adult HIV patients who ever started ART in public health facilities of Debre Markos city were the source population for this study, while complete records of all adult HIV patients in selected public health facilities at Debre Markos city were the study population for this study. Records of all adult HIV patients who ever started ART in selected public health facilities and patients who received/are receiving ART for at least 6 months were included in the study. Meanwhile, records of all adult HIV patients who already developed TB at entry to ART and had an incomplete record were excluded from the study.

### Data source

The data source for this study was an institutional database usually called Smart Care. Healthcare providers manually record patient information on intake and follow-up forms at the time of ART enrollment and during follow-up, respectively. Data clerks also entered patient information from both the intake and follow-up forms into Smart Care.

### Description of the dataset

The dataset included records of all HIV-positive individuals who have ever initiated ART at the respective public health facilities. It contained socio-demographic, clinical, and medication-related information for all HIV patients who had ever started ART in these facilities. In total, 3500 adults have ever started HIV care at DMCSH, and 1892 at Debre Markos Health Center. Therefore, the study focuses on adult HIV patients who have ever initiated ART at these two centers, with a combined total of 5392 individuals who have started ART.

### Study variables

The outcome of interest is the TB status of the patient, which is described as positive (1) or negative (2). Variables that affect the incidence of TB among HIV patients are grouped as sociodemographic, baseline clinical and laboratory characteristics such as nutritional status, and medication-related characteristics, which include type of ART regimen, TPT status, duration on ART, and adherence to ART.

### Operational definitions

Viral load: Suppressed if the viral load is < 50 copies per ml; low-level viremia if the viral load is between 50 and 1000 copies per ml; and high viral load if the viral load is > 1000 copies per ml for clinical intervention [14].

Level of adherence to ART drugs in this study is classified as “good” (≥95% adherence or missing 1 out of 30 doses or missing 2 out of 60 doses). Fair (85–94% adherence or missing 2–4 out of 30 doses or missing 4–9 out of 60 doses), Poor (less than 85% or missing ≥5 doses out of 30 doses or ≥10 doses out of 60 doses) [14].

CD4: severe immune suppression, 200–349 cells/mm³ moderate immune suppression, 350–499 cells/mm³ mild immune suppression, and CD4 counts of 500 cells/mm³ or higher as normal immune function [14].

### Modeling framework

#### Data collection

Data collection began immediately after receiving ethical approval from the Debre Markos University ethical review board. Information was gathered from Debre Markos Health Center and Debre Markos Comprehensive Specialized Hospital. Two data collectors were assigned to extract data from the HIV database at these facilities. The data was initially extracted in Excel format and then converted to CSV format for compatibility with the Python environment.

#### Data Cleaning and Preprocessing

Handling Missing values: Columns such as fluconazole start date, fluconazole end date, and specimen types contained no records and were consequently removed. Meanwhile, columns for address status, WHO clinical stage, months, and ICT status had a combined 1.8% missing values, which were filled using mean and mode imputation.

Data encoding: To enhance data readability, one-hot encoding was applied to categorical variables, including sex, address status, TPT status, viral load status, DSD category, ICT status, follow-up, WHO stage, adherence, nutritional status, and TB incidence.

##### Handling class imbalance

Class imbalance in machine learning refers to a situation where the distribution of instances across different classes is not uniform, leading to a disproportionate representation of one class over another. This imbalance can significantly affect the performance of machine learning models, particularly in classification tasks, as models may become biased towards the majority class and fail to accurately predict the minority class [15]. The dataset contained 4,178 instances (77.5%) in the negative class, while the remaining 1,214 instances (22.5%) were in the positive class. According to the study conducted by Mulugeta et al., if the percentage of data in the minority class is between 20-40%, 1-20%, or less than 1% of the dataset, the class imbalance is considered mild, moderate, or extreme, respectively [16]. Based on this evidence, the dataset exhibited a mild class imbalance, which required the application of techniques to address this issue. Under-sampling, SMOTE, SMOTE+ENN, SMOTE+Tomek, Tomek Links, and ADASYN are all techniques available to handle class imbalance in datasets, particularly in machine learning [17, 18]. As a result, this study compared each technique with the random forest algorithm using accuracy and AUC as evaluation metrics.

#### Feature Selection

Feature selection refers to the process of selecting a subset of the most relevant features (variables) from the original set of features to use in model training. The goal is to reduce the dimensionality of the data, improve model efficiency, and avoid overfitting [19]. Filter methods, including the use of a correlation matrix, are commonly employed in machine learning to select relevant features. In this research, the correlation matrix was utilized as part of the filter method to identify and eliminate highly correlated features, ensuring that only the most informative and non-redundant features were retained for model training. This approach helps to improve model performance and reduce overfitting by focusing on the most relevant variables.

#### Data Split

The technique of splitting a dataset into distinct subsets for testing and training is known as “data splitting” in machine learning. Data is usually divided into two sets: a training set for training the model and a test set for evaluating the model’s performance and ability to generalize to new data. Depending on the size of the dataset and the issue at hand, split ratios can vary, but typically they are 80% for training and 20% for testing [20]. Consequently, this research divided the dataset into 80% (4314) for training and 20% (1078) for testing.

#### Model selection

Model selection in machine learning is the process of choosing the most appropriate algorithm or model for a given problem based on factors like the nature of the data, the task at hand, and performance metrics [17]. In this study, seven supervised classification algorithms, including support vector machine, random forest, decision tree, logistic regression, gradient boosting, K-nearest neighbors, and XGBoost, were selected. After the selection process, each model was trained, and the one with the highest performance metrics was chosen (Figure 1).

**Figure 1:**
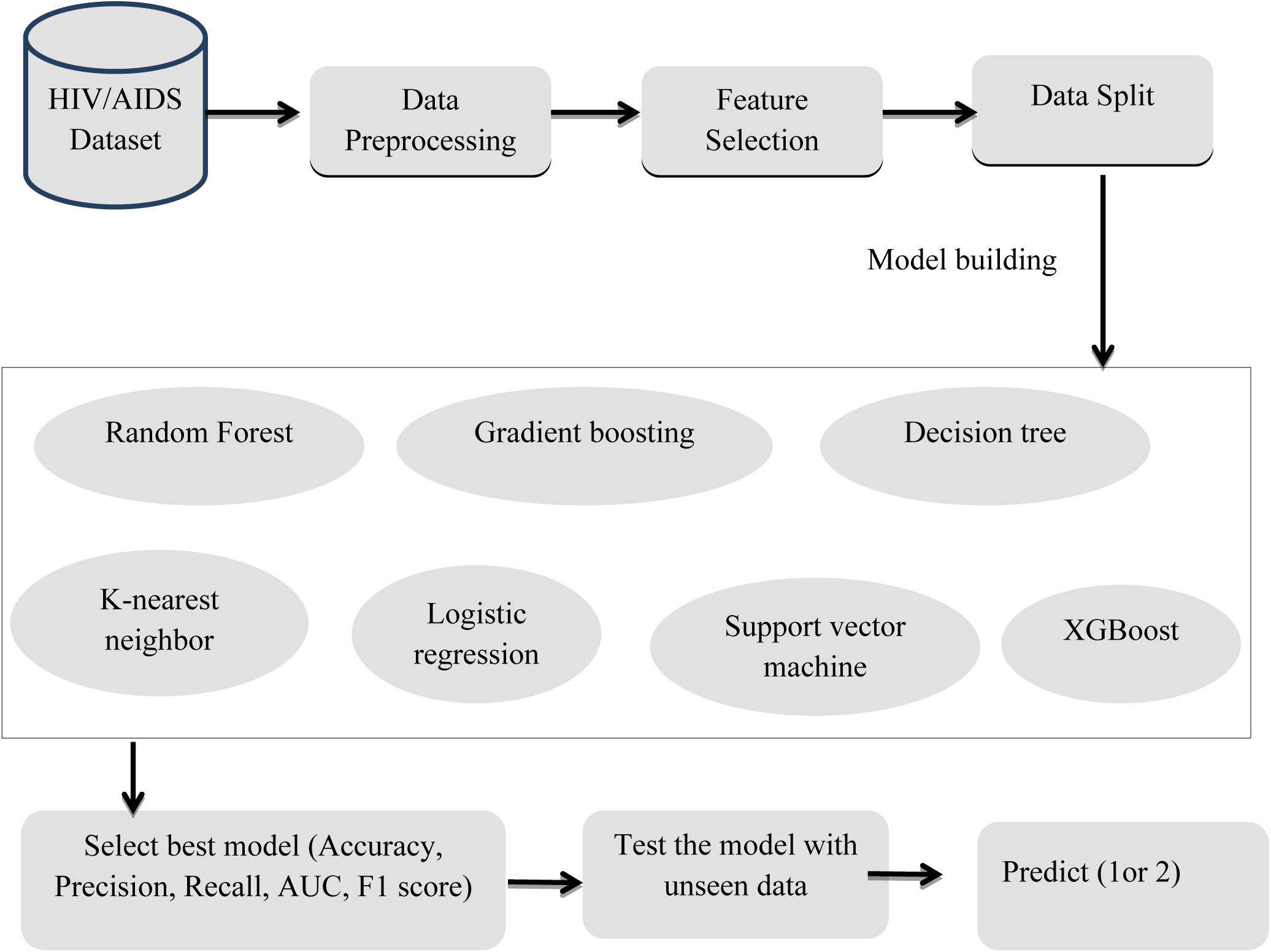
ML modeling framework for predicting incidence of TB among adult HIV patients on ART

**Figure 2:**
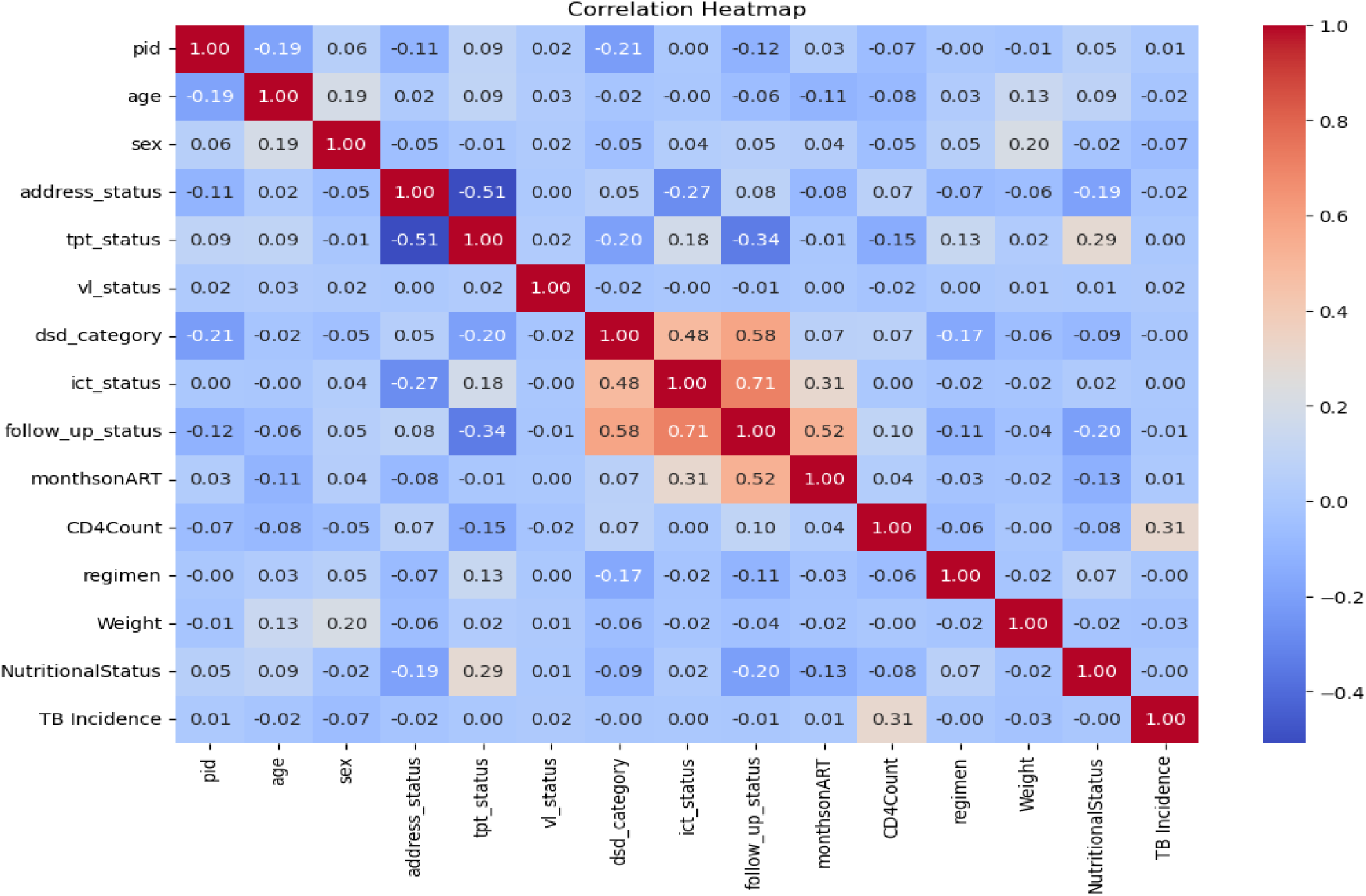
Heat map displaying feature correlation in a study conducted in public health facilities in Debremarkos city, Ethiopia

**Figure 3:**
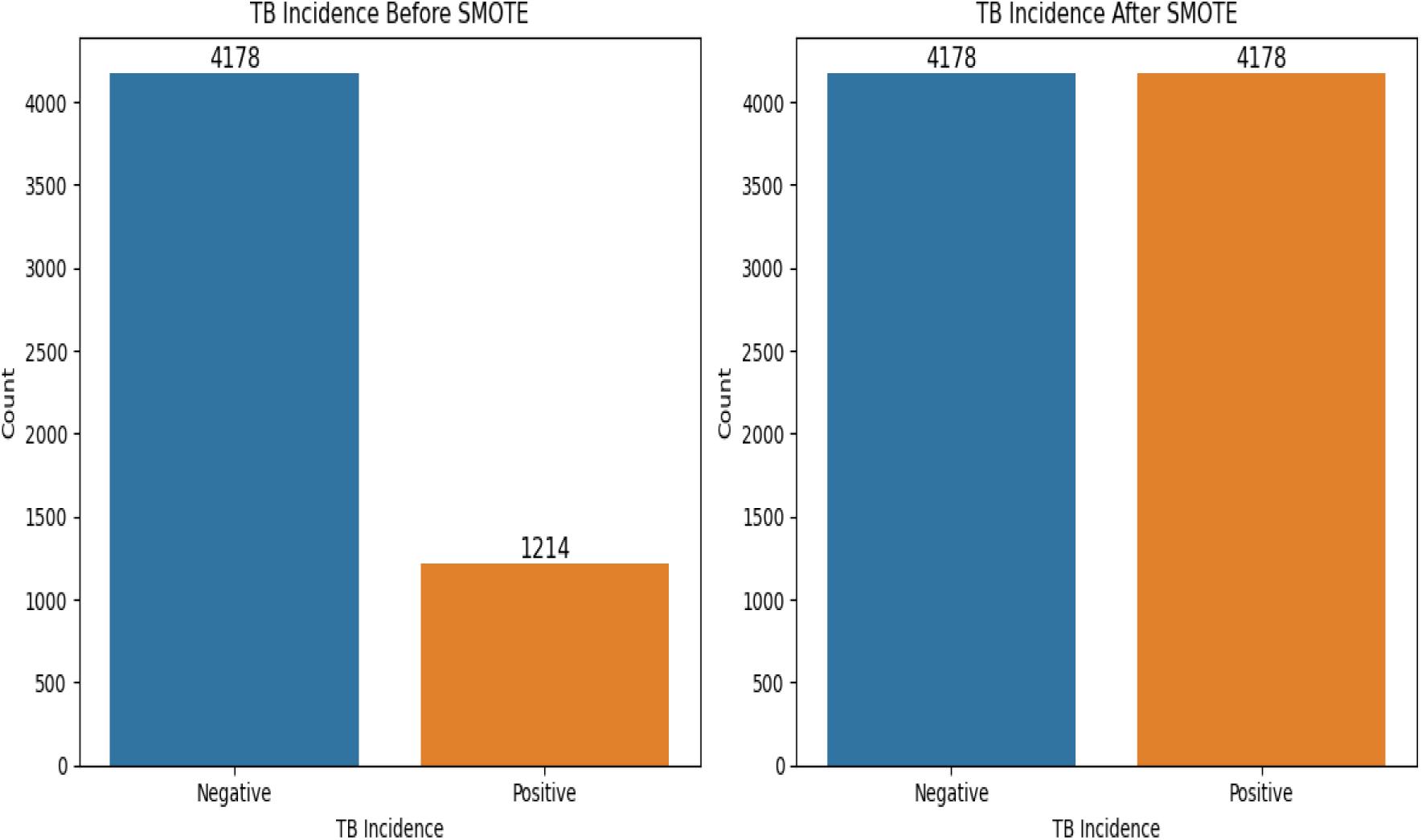
Description of classes before and after applying SMOTE+ENN technique in the adult HIV patients’ dataset from public health facilities of Debre Markos city, 2024

#### Evaluation measures

After model training, the model’s performances were evaluated and compared to each other. The performance of the prediction models was evaluated based on the confusion matrix. This study used precision, sensitivity, specificity, the F1-score, and the area under the receiver operating characteristic (AUC-ROC) to assess the model performance. AUC-ROC is a popular and powerful performance metric for assessing the performance of binary classifiers and is used to evaluate the model’s predictive classification capacity [17]. The matrix is made up of predictions that have been summarized into a total number of correct and incorrect predictions [6].

## Results

### Description of socio-demographic predictors

The study included 5,392 records of adult HIV patients who had ever initiated ART. The average age of the participants was 42 years, with a standard deviation of ±11 years. Among the records, 4,359 (80.8%) were of patients aged 44 years or older, while 1,033 (19.2%) were between the ages of 18 and 43. Of the total records, 3,440 (63.8%) were from female patients, and 1,952 (36.2%) were from male patients. In terms of the patients’ addresses, 3,715 (68.9%) were associated with green records, and 1,677 (31.1%) had yellow records.

### Description of clinical predictors

Out of the total records of adult HIV patients, 4,402 (81.6%) had a normal nutritional status, while 630 (11.7%) were classified as overweight and 360 (6.7%) as undernourished. In terms of WHO clinical stages, 4085 (75.8%) were at stage 1, 549 (10.2%) at stage 2, 538 (10%) at stage 3, and 220 (4%) at stage 4. Regarding CD4 count, 2478 (46%) had counts of ≥500 cells/mm³, 2374 (44%) had counts between 350 and 499 cells/mm³, and 540 (10%) had counts between 200 and 349 cells/mm³. Of the total, 5148 (95.5%) had a suppressed viral load, while 244 (4.5%) had an unsuppressed viral load.

### Description of ART and medication-related predictors

In terms of the regimen type, 98% (5278) of the participants were on first-line drugs, while 2% (114) were on second-line drugs. Regarding the TPT (Tuberculosis Preventive Treatment) status, the records for 51.7% (2781) of participants were categorized as gold, 47.8% (2579) as bronze, and 0.6% (32) as silver.

### Exploratory data analysis

For feature selection, a correlation matrix was used to identify and visualize features that were highly correlated (table 2).

The findings clearly show a class imbalance between the two instances (positive and negative). As a result, this study evaluated several techniques—under-sampling, SMOTE, SMOTE+ENN, SMOTE+Tomek, Tomek Links, and ADASYN—based on accuracy and AUC. Ultimately, the SMOTE+ENN technique was chosen to balance the classes (Table 1).

**Table 1:** Comparison of class imbalance handling techniques for predicting TB incidence among adult HIV patients.

| Technique | Accuracy | ROC-AUC |
| --- | --- | --- |
| Default | 0.753074 | 0.696191 |
| SMOTE | 0.816886 | 0.905226 |
| ADASYN | 0.808295 | 0.896512 |
| Random Under Sampling | 0.645729 | 0.709026 |
| Tomek Links | 0.732873 | 0.731140 |
| SMOTE+ENN | 0.898389 | 0.963591 |
| SMOTE + Tomek | 0.829392 | 0.914808 |

### Model training and evaluation

This study employed 7 machine learning algorithms, using 80% of the dataset for training. Among them, XGBoost performed better than the other algorithms (Table 2).

**Table 2:**
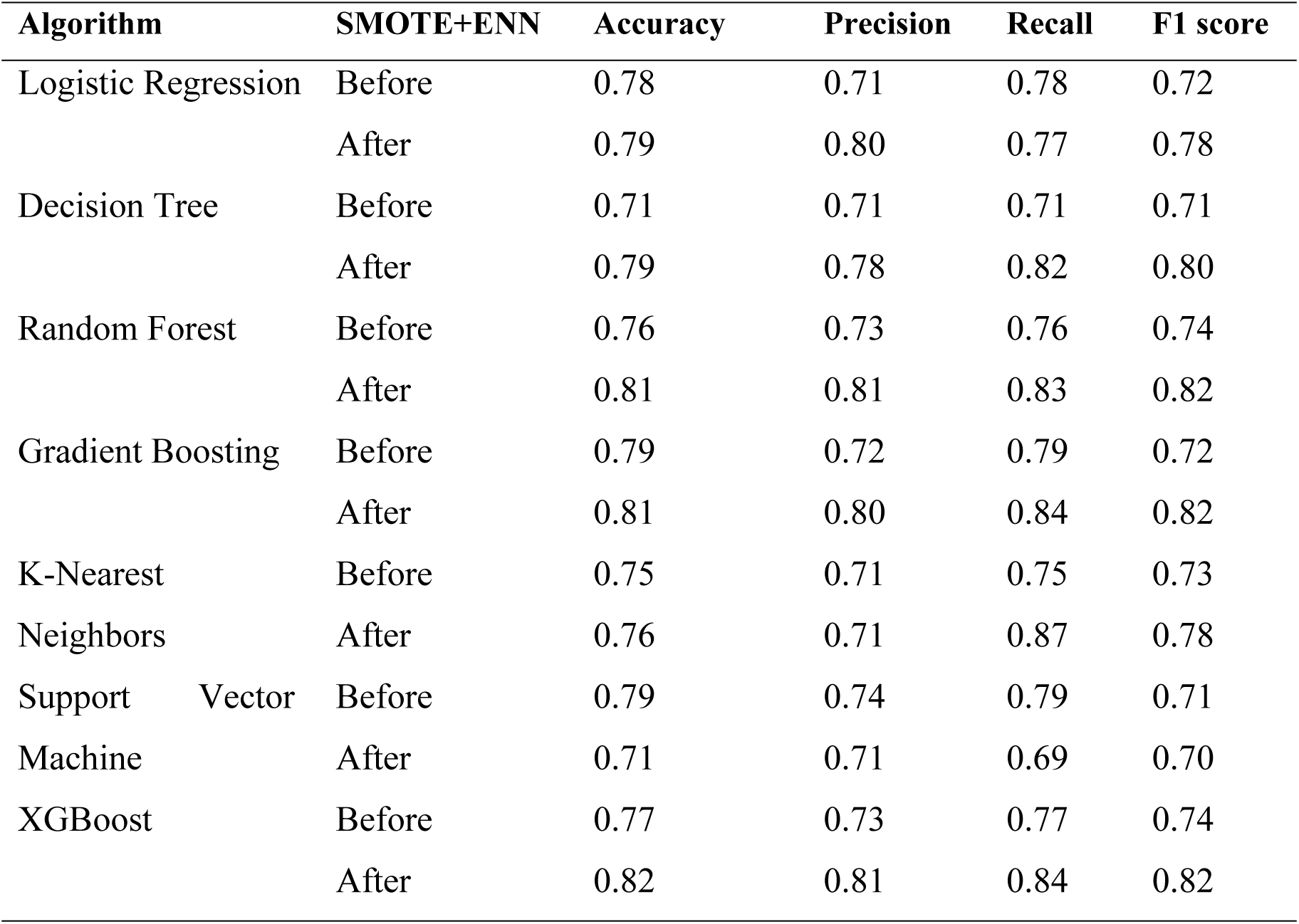
Comparison of algorithms based on accuracy, precision, recall, and F1 score before and after applying SMOTE +ENN using the adult HIV patients’ dataset from public health facilities of Debre Markos City, 2024.

The relationship between false positive and true positive rates is shown in the ROC curve, which states that a predictive model was more accurate for the prediction of TB incidence (Figure 4).

**Figure 4:**
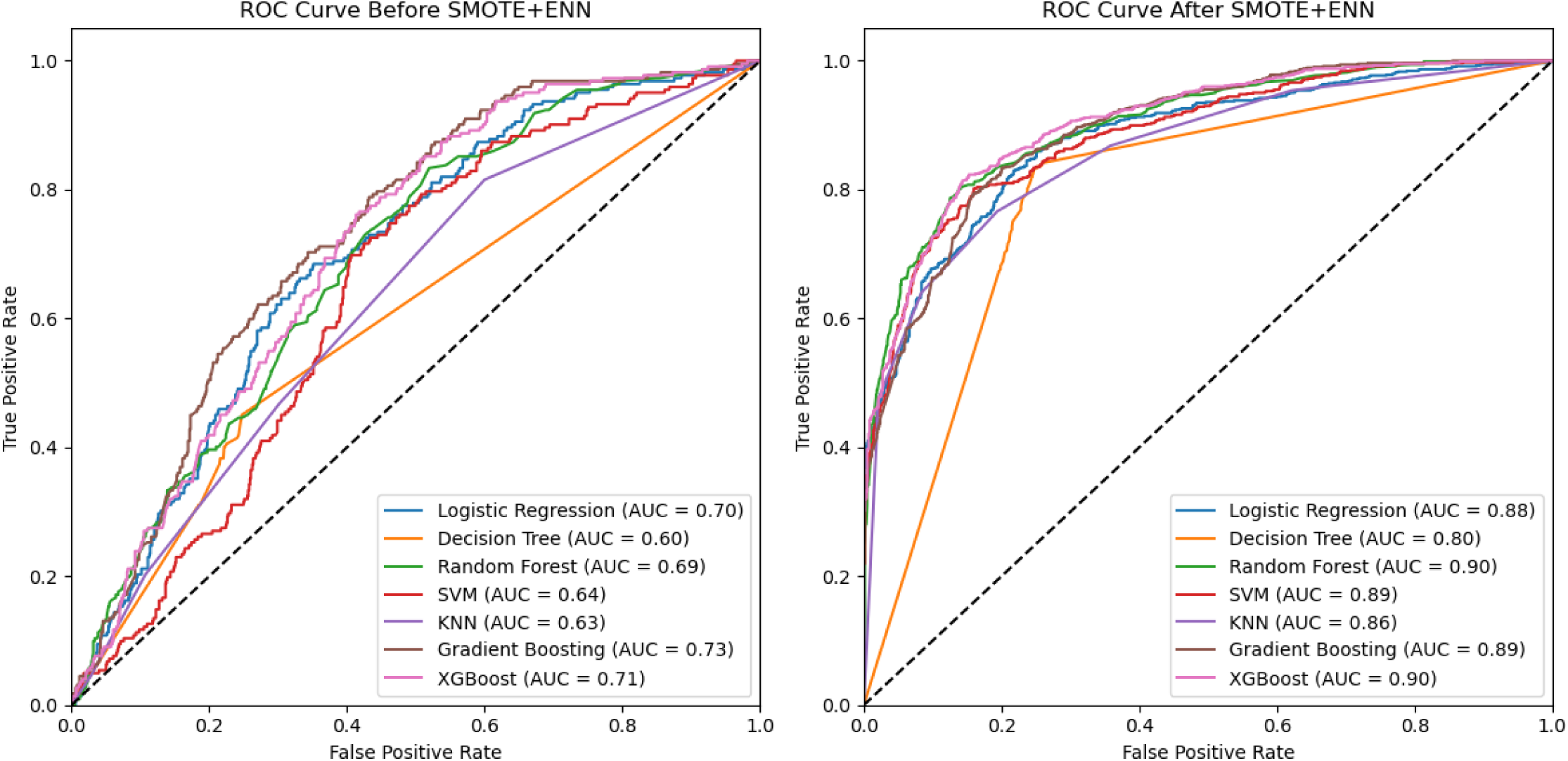
ROC Curve comparing the performance of algorithms using the adult HIV patients’ dataset from public health facilities in Debre Markos City, 2024

### Model testing

To assess the performance of the selected model, it was tested on unseen data, which comprised 1078 instances (20% of the dataset). The model generated a confusion matrix, which included predicted positive vs. actual positive, predicted negative vs. actual negative, and predicted negative vs. actual positive instances (Figure 5).

**Figure 5:**
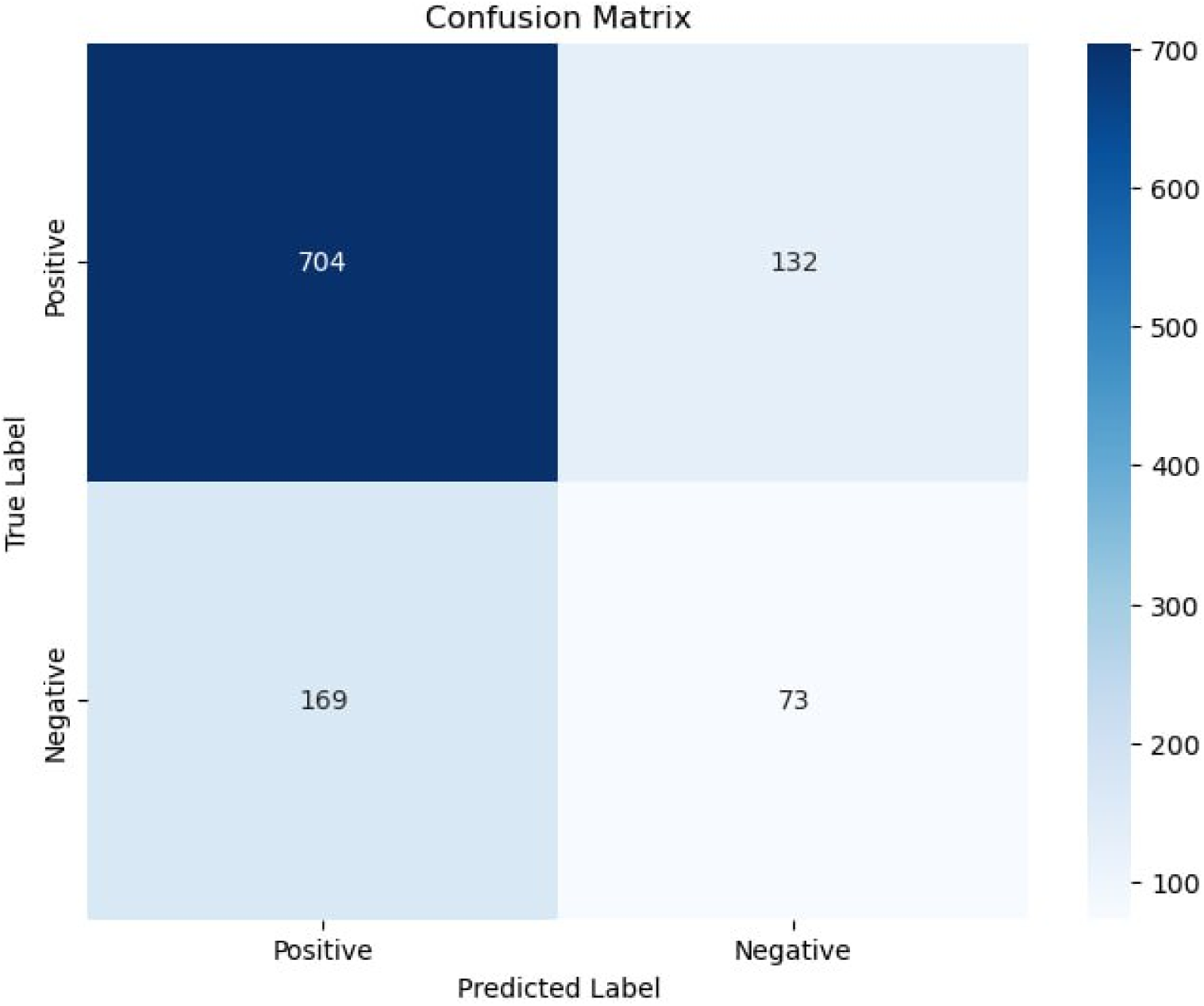
Confusion matrix for XGBoost to predict incidence of TB among adult HIV patient on ART in public health facilities of Debre Markos city, 2024

**Figure 6:**
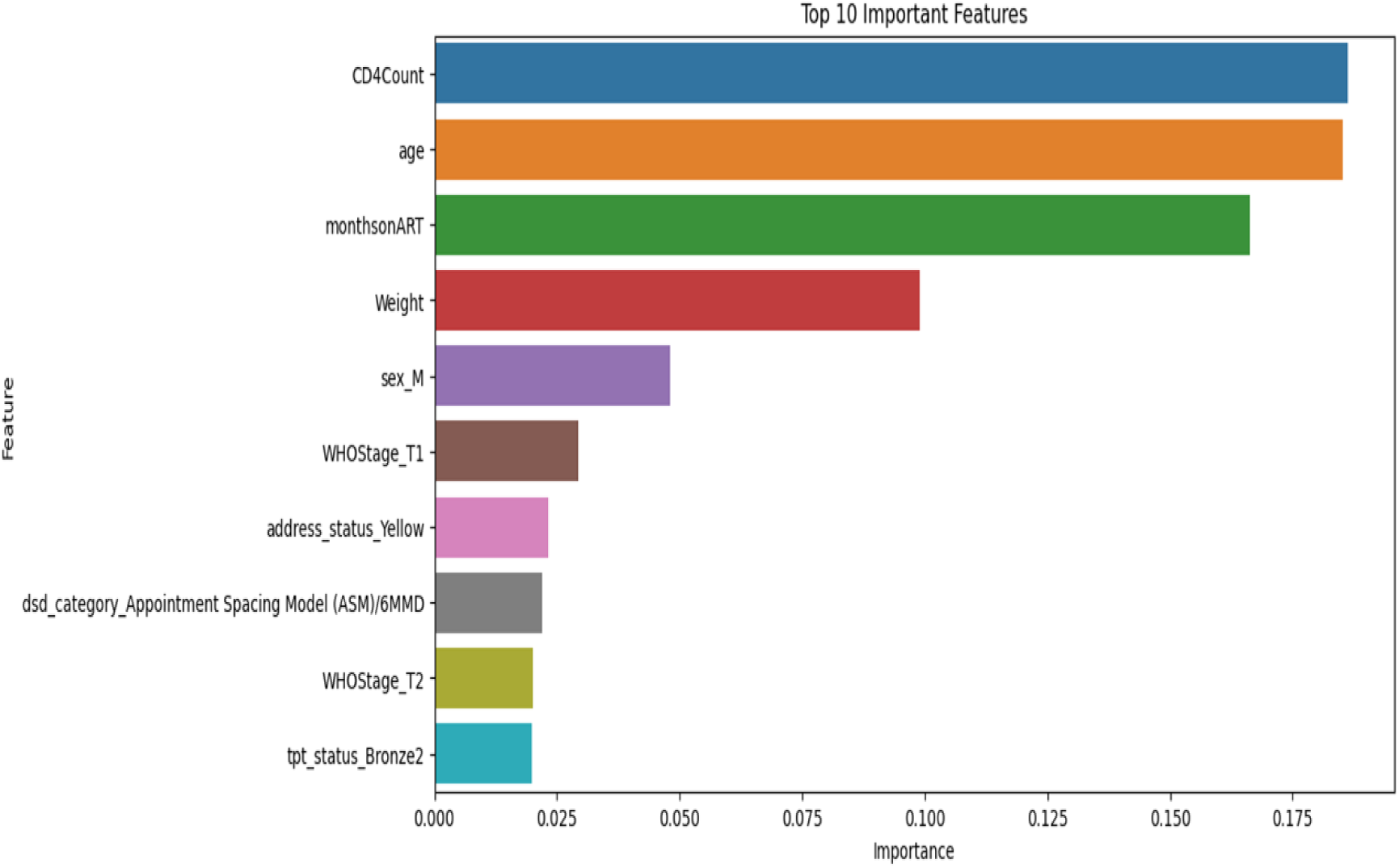
Important features selected by XGBoost feature importance to predict TB incidence among adult HIV patients on ART in public health facilities of Debre Markos, 2024

### Predictors of incidence of TB

This study employed the XGBoost classifier for feature importance selection. Among all the features, CD4 count, age, months on ART, sex, WHO clinical stage, address status, DSD category, and TPT were found to be predictors of incidence TB

## Discussion

Predicting the occurrence of TB and determining its predictors among adult HIV patients at public health institutions in Debre Markos, Ethiopia, was the goal of this study. This was accomplished by evaluating seven machine learning methods for TB prediction among HIV-infected people; XGBoost was found to outperform over other techniques. These results were consistent with research conducted in Taiwan and China, where the XGBoost algorithm was found to be the most successful classifier [13, 21].

The model that was selected for this study, XGBoost, correctly identified 82% of all instances in the training dataset—both positive and negative—as actual positive or actual negative. This outcome nearly aligns with a Chinese study in which 84% of the training dataset’s occurrences were correctly classified by a similar model [21]. Nonetheless, it surpasses a Taiwanese study in which 70.5% of the training dataset was properly classified by the system [13]. Differences in the source population, algorithms included in the training, data splitting techniques, included characteristics, and dataset size may all be responsible for the discrepancies in classification performance.

The XGBoost model in this research correctly classified 84.2% of predicted positives out of all actual positives, achieving a recall (sensitivity) of 84.2%. This performance surpasses the outcome of a Chinese study in which 71% of the true positives were correctly classified by the algorithm [21]. However, the outcome is comparable to a study conducted in Taiwan, where 81.2% of the training dataset’s actual positives were correctly classified as predicted positives [13]. Given that both this study and the Taiwanese study used training datasets that were comparatively bigger than the Chinese study, the discrepancies in the results between these three studies could be the result of disparities in dataset sizes.

The AUC in this study is 90%, indicating a 90% probability that the model will correctly rank a randomly selected positive instance higher than a randomly selected negative one. This result is slightly higher than a study conducted in Taiwan, where the model demonstrated an 86.2% probability of correctly distinguishing a randomly chosen positive instance from a randomly chosen negative one [13]. This difference could be explained by variations in the dataset size. The dataset used in this study was approximately twice as large as the one used in the Taiwanese research.

CD4 count, age, duration on ART, sex, WHO clinical stage, address status, DSD category, and TPT were identified as predictors of tuberculosis incidence among adult HIV patients in public health facilities in Debre Markos city. Machine learning methods used for predicting tuberculosis risk factors highlighted age and length of ART as significant risk factors for TB [22]. Additional research conducted in Ethiopia using statistical modeling found that a low CD4 count, not taking IPT prophylaxis, and being in WHO stage III or IV at baseline were associated with an increased risk of tuberculosis [23]. Similarly, a study in Tanzania identified male sex, lower CD4 counts, and advanced WHO disease stages as predictors of TB incidence [24].

### Conclusion

This study developed and validated machine learning models to predict TB incidence in adult HIV/AIDS patients who are at ART centers. In our study, support vector machine, random forest, decision tree, logistic regression, gradient boosting, K-nearest neighbors, and XGBoost models have been tested. The XGBoost models have provided strong predictive performance. The current study demonstrated that machine learning-based prediction models are promising tools in accurately predicting tuberculosis incidence in adult HIV/AIDS patients. Further external validation and the inclusion of additional factors are needed to enhance the model’s robustness and generalizability.

### Limitations of the research

A limitation of this research is that the data was sourced from secondary datasets originally collected for different purposes, which resulted in the absence of certain features, such as BMI. Additionally, the available records contained 1.8% missing values, which were imputed, and this could also have influenced the study’s outcomes.

## Data Availability

Data is available from PI and will be avail based on request

## Ethical Approval

Ethical approval was granted by the Institutional Research Ethics Review Committee of Debre Markos University. A letter of cooperation was received from the Amhara Public Health Institute, Debre Markos Branch. Additionally, permission was obtained from both Debre Markos Comprehensive Specialized Hospital and Debre Markos Health Center.

## CRediT authorship contribution description

***Desalegn Meseret Tadele:*** *Writing original draft, Methodology, Investigation, Formal analysis, Conceptualization. **Getaye Tizazu Biwota:** Supervision, Methodology, review and editing, and manuscript preparation. **Lijalem Megbaru Enyew :** Supervision, Methodology, review and editing. **Maru Meseret Tadele:** Resources, Methodology and Formal analysis. **Gizaw Hailiye Teferi:** Supervision and formal analysis and validation*.

## Funding

*This research did not receive any specific grant from funding agencies in the public, commercial, or not-for-profit sectors.*

## Conflicts of interest

*No conflict interest claim among authors*

## Acknowledgments

None

## Abbreviations and acronyms

AIDS: Acquired Immune Deficiency syndrome
ALWHIV: Adults Living with HIV
ART: Antiretroviral Therapy
AUC: Area Under the Curve
BMI: Body Mass Index
CD4: Cluster of differentiation 4
CDC: Center for Disease Control
CPT: Cotrimoxazole Prophylactic Therapy
DMCSH: Debre Markos Comprehensive Specialized Hospital
DSD: Differentiated Service Delivery
ENN: Edited Nearest Neighbor
HER: Electronic Health Record
HIV: Human Immune Virus
ICT: Index Case Test
IPT: Isoniazid Preventive Therapy
ML: Machine Learning
MoH: Ministry of Health
OIs: Opportunistic Infections
ROC: Receiver Operating Characteristic
SMOTE: Synthetic Minority Over Sampling Technique
TB: Tuberculosis
TPT: Tuberculosis Preventive Therapy
WHO: World Health Organization

## Notes

### Competing Interest Statement

There are not any competing interests

### Funding Statement

The author(s) received no specific funding for this work.

### Author Declarations

Ethical approval was granted by the Institutional Research Ethics Review Committee of Debre Markos University

